# Bone changes assessed with High-Resolution peripheral Quantitative Computed Tomography (HR-pQCT) in early inflammatory arthritis: a 12-month cohort study

**DOI:** 10.1101/2019.12.20.19015537

**Authors:** Scott C. Brunet, Stephanie Finzel, Klaus Engelke, Steven K. Boyd, Cheryl Barnabe, Sarah L. Manske

## Abstract

**Objectives:** We sought to determine the sensitivity of high resolution peripheral quantitative computed tomography (HR-pQCT) to detect change and identify erosions in comparison with conventional radiography (CR) in early inflammatory arthritis patients. We also explored which prognostic factors contribute to bone damage assessed by HR-pQCT in the first year of diagnosis.

**Methods:** 46 patients with arthritic symptoms less than one year, and a clinical diagnosis of inflammatory arthritis were prospectively imaged at baseline and 12-months. HR-pQCT scans of the 2^nd^ and 3^rd^ MCP joints and CR of the hands and feet were performed. Joint space width (JSW), total bone mineral density (Tt.BMD), erosion presence and volume were assessed with HR-pQCT. Scan-rescan precision was assessed to define an individual-level least significant change (LSC) criterion. Regression analyses explored prognostic factors for bone damage progression.

**Results:** We observed no significant group-level changes in JSW, Tt.BMD or erosion volume. 20% or fewer joints demonstrated individual-level changes greater than the LSC criterion for mean JSW, Tt.BMD and erosion volume. HR-pQCT detected more erosions than CR in the 2^nd^ and 3^rd^ MCP. Increased symptom duration at diagnosis was associated (p < 0.10) with lower JSW minimum and higher JSW standard deviation.

**Conclusions:** We have demonstrated stability in erosion, bone density and JSW over 12-months in most patients receiving treatment for inflammatory arthritis using a LSC criterion, although patients with longer symptom duration prior to treatment initiation had demonstrable negative effects on JSW estimates. HR-pQCT captures bone damage and progression undetectable by CR in the imaged joints.

## INTRODUCTION

Early diagnosis and tight monitoring of disease progression are important clinical targets in patients with inflammatory arthritis to optimize treatment response and prevent future damage.[1-3] Conventional radiographs (CR) are traditionally used to assess erosive disease and joint space loss, however may not be sufficiently sensitive to detect changes early in the disease course.

High-resolution peripheral quantitative computed tomography (HR-pQCT) is a highly sensitive bone imaging tool, capable of capturing 3D volumetric bone images at 61 or 82 µm nominal isotropic resolution.[4,5] Quantitative measurement techniques have been developed and validated to assess metacarpophalangeal (MCP) joint space width, erosion number and size as well as periarticular bone density and microarchitecture.[6-9] We previously found that HR-pQCT-assessed joint space width outcomes are associated with van der Heijde-modified total Sharp scores (vdHSS) from conventional radiography in patients with advanced disease.[10] These studies provide a strong rationale to investigate the use of HR-pQCT in early inflammatory arthritis patients. The initial disease course in RA is highly heterogenous;[11] predicting who may benefit from aggressive therapy is important to reduce medical risk and cost, as well as preserve function and productivity.[12] While HR-pQCT has been used to demonstrate repair effects at the group level,[13-16] to our knowledge, whether there is utility in monitoring progression with HR-pQCT in the early course of inflammatory arthritis has not been determined. Further, whether established risk factors for aggressive disease, including seropositivity, number of swollen joints and radiographic evidence of erosions [17-20] are also associated with progression of outcomes measured by HR-pQCT is not known.

The purposes of this study were two-fold. First, we sought to demonstrate the relative ability of HR-pQCT to identify erosions in comparison with CR. Second, we explored which factors typically associated with aggressive RA, were associated with baseline bone damage and progression assessed by HR-pQCT, in patients with a recent (< 1 month) diagnosis of early inflammatory arthritis.

## METHODS

### Patients

We recruited 46 patients with early inflammatory arthritis (EIA) from the University of Calgary Division of Rheumatology specialty EIA clinic between 2011 and 2014. Eligible patients had experienced inflammatory arthritis symptoms for less than one year, had a clinical diagnosis of early rheumatoid arthritis (RA) or undifferentiated arthritis by a rheumatologist, and were beginning disease-modifying therapy according to standard clinical practice. All patients provided written informed consent prior to study participation. Approval for all procedures was obtained by the Conjoint Health Research Ethics Board at the University of Calgary (REB 15-0582). Study visits were conducted at baseline and 12-months. Patients were not involved in the design, conduct, reporting, or dissemination plans of our research.

### Demographic and Clinical Outcomes

Demographic information (age, sex, BMI, handedness, occupation, dates of symptom onset and diagnosis) were recorded. Patients were classified according to the 2010 ACR/EULAR Classification for RA [21]. Laboratory diagnostics comprised rheumatoid factor (RF), anti-cyclic citrullinated peptide antibody (ACPA), C-reactive protein (CRP) and erythrocyte sedimentation rate (ESR). Disease activity assessment included tender and swollen joint counts, physician and patient global scores, duration of morning stiffness, Health Assessment Questionnaire (HAQ) scores [22] and 28-joint Disease Activity Score (DAS28).[23] Remission status (DAS28 < 2.6)[24] was also recorded as well as treatments received.

### Conventional Radiographs

Routine conventional plain film radiographs of the hands, wrist and feet were performed. All CR were scored for erosions, joint space narrowing (JSN) and subluxation using the vdHSS by a single experienced reader from Imaging Rheumatology International (Meersan, Netherlands), blinded to all other patient data.[25] Radiographic progression was defined as a single unit increase in vdHSS score.[26]

### HR-pQCT Image Acquisition

HR-pQCT scans were performed on the dominant 2^nd^ and 3^rd^ MCP joints secured in a custom positioning device (XtremeCT, Scanco Medical, Brüttisellen, Switzerland).[27] We obtained a scout view (coronal plane x-ray) to identify the reference line. HR-pQCT scans are acquired in 110 slice increments (“stacks”). For the first 13 baseline scans, the reference line was placed at the distal surface of the 3^rd^ proximal phalangeal bone covering 9 mm distal and 9 mm proximal to the line for a total of 1.8 mm (2 stacks = 220 slices). For the remaining patients, to comply with the Study grouP for eXtreme Computed Tomography in Rheumatoid Arthritis (SPECTRA) recommendations for standard positioning which were agreed to shortly after study initiation [28] and to ensure the joint space was captured by a single acquisition, the reference line was placed on the distal surface of the 2^nd^ or 3^rd^ metacarpal, whichever was more distal, and the scan region of interest included 9.02 mm distal and 18.04 mm proximal to the line for a total of 2.7 cm (3 stacks or 330 slices). Images with a nominal isotropic resolution of 82.0 µm were acquired using the manufacturer’s standard settings (60 kVp, 1000 µA, 100 ms integration time).

### HR-pQCT Image Processing

HR-pQCT image processing details are provided in Appendix 1. Briefly, we assessed total bone mineral density (Tt.BMD, mg/cm^3^) from grey-scale images. Volumetric joint space was quantified using an algorithm developed by consensus from the SPECTRA collaboration.[29] 3D JSW including mean (JSW.Mean, mm), maximum (JSW.Max, mm), minimum (JSW.Min, mm), standard deviation (JSW.SD, mm), asymmetry (defined as JSW.Asymm = ratio of JSW.Max/JSW.Min, [1]) as well as volume (JSV, mm^3^) were calculated. Erosion presence was identified by a rheumatologist with extensive HR-pQCT expertise (SF). Semi-automated assessment of erosion volume was performed using the Medical Image Analysis Framework (University of Erlangen, Erlangen Germany).[9]

### Precision Analyses

Short-term *in vivo* precision data were obtained by performing a second scan on the same day with repositioning.[30] In addition, intra-operator precision was assessed for the erosion analysis. Precision of JSW outcomes was previously reported in this cohort. [29] Here precision of erosion volume and Tt.BMD are reported.

### Statistical Analyses

For all demographic, clinical and HR-pQCT outcomes, results are reported as mean and standard deviation (SD) unless otherwise indicated. For HR-pQCT precision, we calculated the precision error as the root-mean square standard deviation (RMSSD) and the root-mean square coefficient of variation (RMSCV).[31] For erosion volume, precision was calculated separately for erosions less than 10 mm^3^ and erosions greater than 10 mm^3^ for comparison with previously published values.[9] In addition, the distribution of the difference between scan-rescan values were compared to determine whether they met the criteria required to compute the least-significant change (LSC) at the 95% confidence level (i.e., 2.77 x RMSSD).[32]

Paired Welch’s t-tests were used to examine changes over time at the group level, and tested against an alpha level of 0.05. For paired differences that were not normally distributed, a paired Wilcoxon signed-rank test was used. Beeswarm plots were used to examine the heterogeneity of the data.[33] To determine whether an individual change exceeded measurement error at a 95% confidence level, the changes in mean JSW, JSV, TtBMD and erosion volume were evaluated against the LSC, as described above.[32] Using the LSC criteria, patients were classified into one of three groups for each outcome measure: improvement, stable, or progression. Based on these criteria, Fisher’s exact test was used to determine whether patients with a diagnosis of RA differed from those with self-limited or undifferentiated arthritis, as well to compare patients with and without: high disease activity at baseline (DAS28 > 5.1),[34] progression to biologic therapy, and remission (DAS28 < 2.6) [24] at follow-up.

To understand whether HR-pQCT outcomes are associated with known prognostic factors and disease activity markers, we performed exploratory analyses using simple linear regressions to examine the relationship between prognostic factors and joint space, erosion volume and bone density. Regressions were performed for the 2^nd^ and 3^rd^ MCPs independently and tested against an alpha value of 0.1 due to the exploratory nature of the analyses. Linear regression relationships with an adjusted r-squared greater than zero and statistical significance across both joints were investigated further. Similarly, binomial logistic regression was used to evaluate the relationship between prognostic factors and erosion presence and tested against an alpha value of 0.1. These analyses were performed for both baseline HR-pQCT outcomes and change in HR-pQCT outcomes. All statistical analyses were performed using R (v3.5.3), RStudio (v1.1.463)[35] and RMarkdown.[36,37]

## RESULTS

### Demographics and Clinical Characteristics

Of the 46 patients recruited, one withdrew from the study prior to follow-up and two were lost to follow-up. Table 1 shows characteristics of the 43 patients who attended both study visits. At baseline, 33 (77%) patients met the 2010 American College of Rheumatology criteria for RA [21] and 10 (23%) were thus assessed to have undifferentiated arthritis. At 12-month follow-up, several patients were reclassified, resulting in a final distribution of 37 (86%) with RA, 2 (5%) still being assessed as having undifferentiated arthritis, 1 (2%) with Adult Onset Still’s, 1 (2%) with psoriatic arthritis, and 2 (5%) with self-limited arthritis. At baseline, 4 (10%) of patients had been exposed to a conventional disease-modifying anti-rheumatic drug (cDMARD), while 38 (88%) utilized a cDMARD over the 12-month follow-up. Eleven (26%) of patients progressed to requiring a biologic DMARD (bDMARD) over the 12-month follow-up. 38 (97%) of patients had active disease (DAS28 ≥ 2.6) at baseline, while 13 (35%) had active disease at follow-up, with mean reductions in the tender and swollen joint counts over one year (Table 1).

**Table 1.** Characteristics of cohort. Mean (SD) for continuous variables. P-values represent significance levels for paired t-tests (*) or Wicoxon paired signed-rank test (#) comparing baseline to 12-month follow-up. BMI = body mass index, DAS-28 = disease activity score with 28 joint count, HAQ = Health Assessement Questionnaire, AM = morning, cDMARD = conventional disease modifying anti-rheumatic drug, bDMARD = biologic DMARD, RF = rheumatoid factor, ACPA = anti-cyclic citrullinated peptide antibody, ESR = erythrocyte sedimentation rate, CRP = c-reactive protein. (n = 43).

|  | Baseline | 12-month Follow-up | p-value |
| --- | --- | --- | --- |
| Age (years) | 46.4 (14.8) | - |  |
| Sex (female:male) | 29:14 | - |  |
| BMI (kg/m<sup>2</sup>) | 27.5 (6.5) | - |  |
| Symptom Duration (days) | 200 (118) | - |  |
| Diagnosis Duration (days) | 33 (47) | - |  |
| **Clinical** |  |  |  |
| DAS-28 | 5.3 (1.2) | 2.3 (1.1) | < 0.001* |
| 28 Swollen Joint Count | 10 (7) | 1 (3) | < 0.001* |
| 28 Tender Joint Count | 11 (8) | 1 (3) | < 0.001# |
| HAQ (0-3) | 1.2 (0.7) | 0.5 (0.5) | < 0.001* |
| Patient Global Score (0 -100) | 55 (23) | 24 (19) | < 0.001* |
| Duration of AM stiffness (min) | 141 (140) | 34 (49) | < 0.001# |
| Remission (DAS28 < 2.6) | 1/39 (3%) | 13/37 (35%) |  |
| Use of cDMARDs | 4/42 (10%) | 38/43 (88%) |  |
| Use of steroids | 13/42 (31%) | 18/43 (42%) |  |
| Use of bDMARDs | 0 | 11/43 (26%) |  |
| **Serological** |  |  |  |
| RF+ | 19/42 (45%) | - |  |
| ACPA+ | 28/40 (70%) | - |  |
| ESR (mm/hr, normal < 20) | 24.7 (18.8) | 11.0 (10.0) | < 0.001# |
| CRP (mg/L, normal < 8) | 22.0 (47.5) | 5.1 (11.0) | < 0.001# |

### Precision

Seventeen joints from 13 patients were used for precision analysis as they had at least one erosion, as well as two scans without motion artifacts acquired on the same day. Intra-rater precision of erosion volume determined as RMSSD was 1.89 mm^3^ for total erosion volume per joint, and 2.52 mm^3^ when evaluating by individual erosion volume. RMSSD for erosions > 10 mm^3^ was 3.49 mm^3^ and for erosions < 10 mm^3^ was 1.25 mm^3^ (Table 2).

**Table 2.** Precision errors calculated for scan-rescan results with repositioning. Intra-rater precision is also presented for erosion volume. JSW precision was previously reported in this cohort in [29]. SD = standard deviation, AS = asymmetry. n = 42 joints for JSW, 30 for Tt.BMD and 17 for Erosion Volume. LSC values are noted as missing (-) when the measurements failed to reach the criteria to calculate the LSC at the 95% confidence interval.

|  | Scan-Rescan |  |  | Intra-Rater |  |  |
| --- | --- | --- | --- | --- | --- | --- |
|  | RMSSD | RMSCV<br>(%) | LSC | RMSSD | RMSCV<br>(%) | LSC |
| JSW |  |  |  |  |  |  |
| Mean (mm) | 0.06 | 3.6 | 0.18 |  |  |  |
| Volume (mm<sup>3</sup>) | 4.26 | 4.3 | 11.8 |  |  |  |
| Minimum (mm) | 0.25 | 26.2 | - |  |  |  |
| Maximum (mm) | 0.09 | 3.1 | - |  |  |  |
| SD (mm) | 0.04 | 11.1 | - |  |  |  |
| AS (1) | 2.1 | 53.7 | - |  |  |  |
| Tt.BMD (mg HA/cm<sup>3</sup>) | 11 | 3.6 | 30 |  |  |  |
| Erosion Volume |  |  |  |  |  |  |
| Total Erosion Volume (mm<sup>3</sup>) | 0.72 | 4.1 | 2.0 | 1.89 | 10.1 | - |
| Individual Erosion (mm<sup>3</sup>) | 1.04 | 7.6 | - | 2.52 | 15.9 | - |
| Erosions < 10 mm<sup>3</sup> (mm<sup>3</sup>) | 0.63 | 18.7 | - | 1.25 | 38.4 | - |
| Erosions > 10 mm<sup>3</sup> (mm<sup>3</sup>) | 1.72 | 4.1 | - | 3.49 | 11.2 | - |

Analysis of scan-rescan precision revealed smaller precision errors; RMSSD was 0.72 mm^3^ for total erosion volume per joint and 1.04 mm^3^ when evaluating by individual erosion volume. RMSSD for erosions > 10 mm^3^ was 1.72 mm^3^ and for erosions < 10 mm^3^ was 0.63 mm^3^ (Table 2). RMSSD for Tt.BMD was 11 mgHA/cm^3^. LSC are reported for those variables that met the required assumptions (mean JSW, JSV, total erosion volume, Tt.BMD, Table 2).

### Relationship between Prognostic Factors and Baseline JSW, TtBMD and Erosion Volume

For quantitative analyses, 4 joints were excluded from baseline JSW, Tt.BMD and erosion volume analysis due to motion artifact or segmentation errors. Males had greater JSV (52 mm^3^ in 2^nd^ MCP), mean JSW (0.3 mm in 2^nd^ MCP) and max JSW (0.1 mm in 2^nd^ MCP) than females in both 2^nd^ and 3^rd^ MCPs at baseline (p < 0.005, adjusted R^2^ varying from 0.19 to 0.69). Increased symptom duration at diagnosis was significantly associated (p < 0.10) with lower minimum JSW (adjusted R^2^ = 0.12 and 0.05) suggesting that for every 100 days of symptoms, there is a 0.01 mm decrease in minimum JSW. Similarly, symptom duration was associated with increased SD of JSW (adjusted R^2^ = 0.06 and 0.24) for 2^nd^ and 3^rd^ MCPs at baseline, suggesting that for every 100 days of symptoms, there is a 0.002 increase in SD. We found no significant associations between baseline age, duration of time between diagnosis and scan, BMI, seropositivity, HAQ score, patient global score, morning stiffness duration, exposure to steroids, or DAS28 score and JSW, Tt.BMD or erosion volume outcomes.

### HR-pQCT Comparison with Conventional Radiography

Seventeen (40%) of patients had erosions identified by HR-pQCT in the 2^nd^ or 3^rd^ MCP at baseline. Two individuals (4%) with erosions at the 2^nd^ or 3^rd^ MCP were identified by CR, but HR-pQCT allowed identification of erosions in an additional 16 patients (38%) with HR-pQCT. Although HR-pQCT was more sensitive to identifying erosions at these joints, there were 11 patients (26%) with erosions captured in additional joints by CR included in the vdHSS scoring system (Figure 1). In the participant with the highest vdHSS, HR-pQCT revealed a “pseudoerosion” (i.e., no cortical break) in the 2^nd^ MCP, but no erosions that met the SPECTRA erosion definition in the 2^nd^ or 3^rd^ MCP (Figure 2).

**Figure 1.**
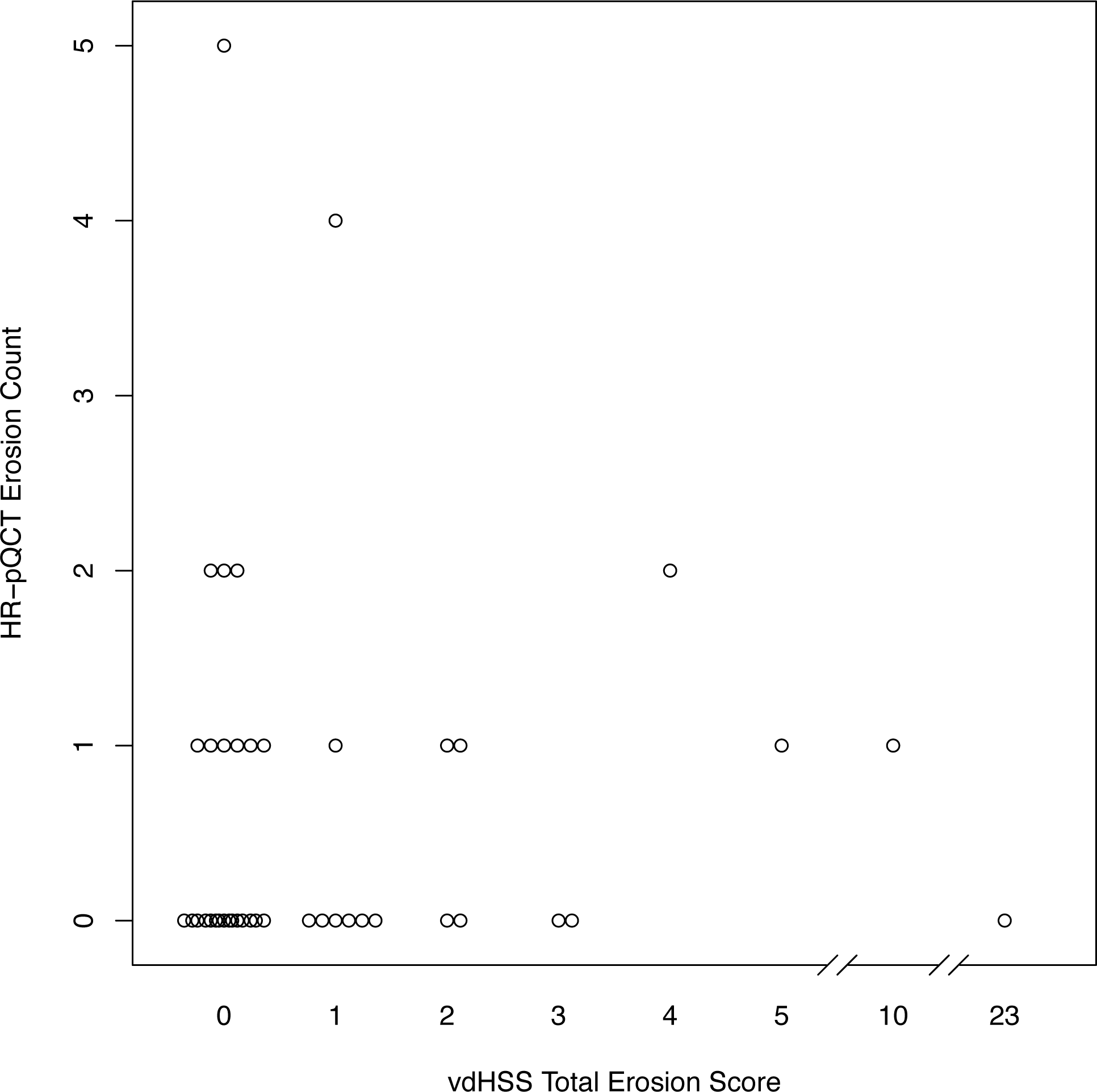
Erosion count by HR-pQCT in MCP2 and 3 compared with vdHSS Total Erosion scores from CR at baseline. (n = 41)

**Figure 2.**
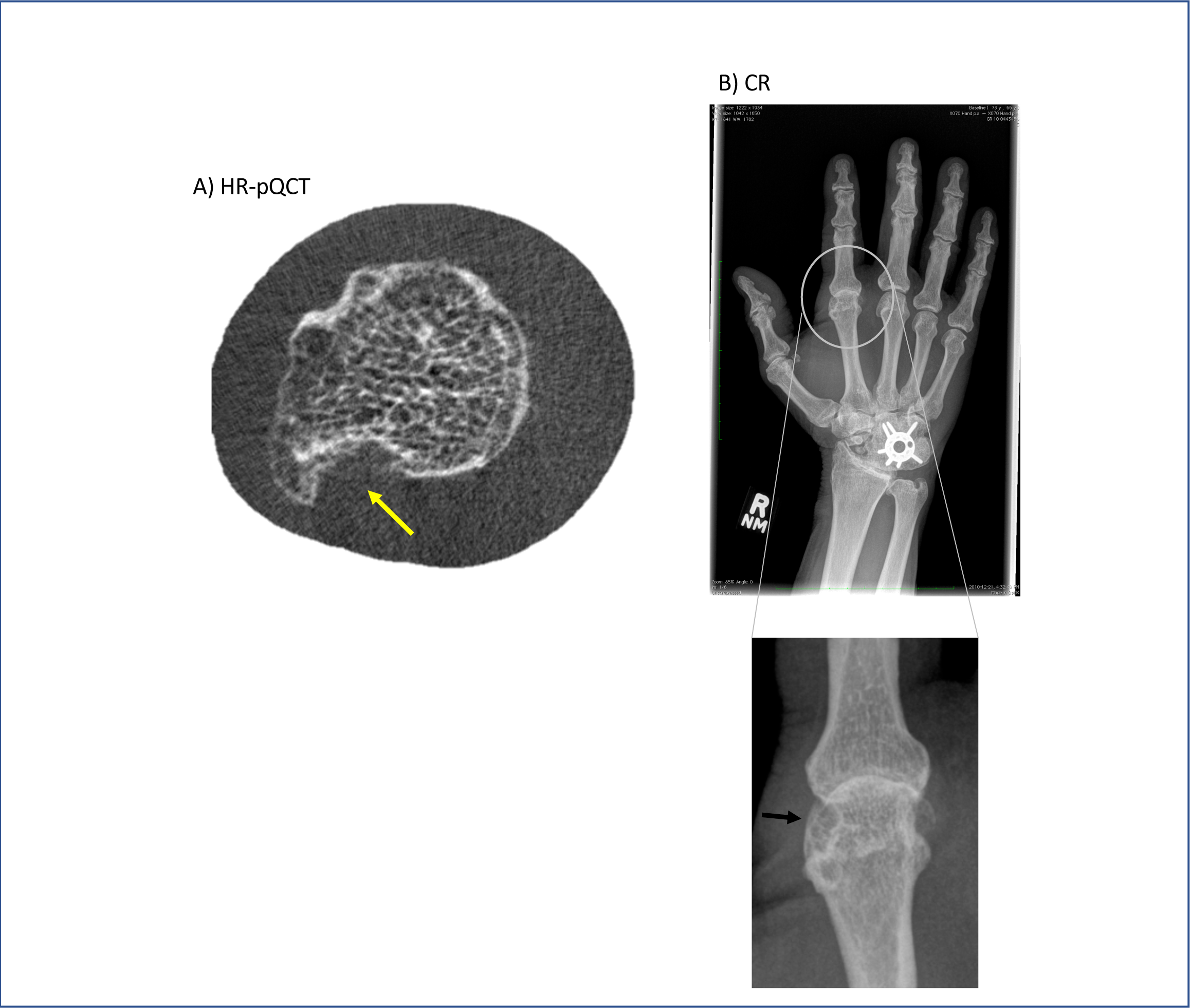
A) Pseudo-erosion (yellow arrow) identified in an axial HR-pQCT of the 2^nd^ MCP.B) Corresponding conventional radiograph (CR) with the 2^nd^ MCP erosion indicated with a black arrow.

Minimum JSW by HR-pQCT was not associated with the vdHSS total JSN score (data not shown). One participant had a non-zero vdHSS JSN score in the 2^nd^ or 3^rd^ MCPs; thus a direct comparison between JSW and vdHSS joint score at these joints could not be made.

### Longitudinal Changes in JSW, Tt.BMD, Erosion Number and Erosion Volume by HR-pQCT

Examination of raw images found two patients who developed erosions by 12-month follow-up. After image registration, the appearance of new erosions was less convincing (Figure 3). The overall number of erosions increased from 28 to 36 between baseline and 12-month follow-up.

**Figure 3.**
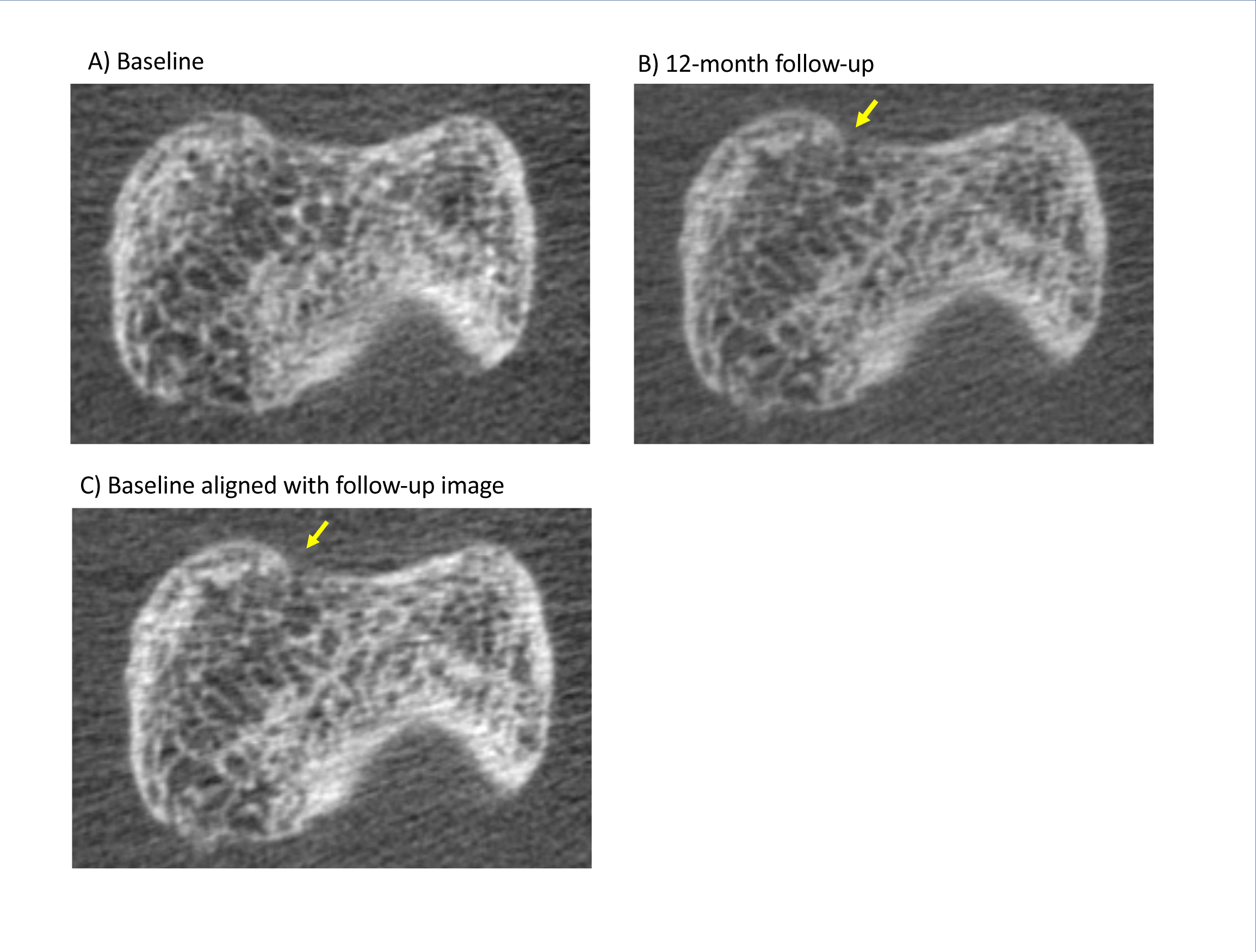
Axial HR-pQCT images of the 2^nd^ phalangeal base demonstrating an apparent new erosion development over the 12-month follow-up. Image registration and transformation of the baseline image to align with the 12-month follow-up image suggests that the observation of a new erosion was likely artifactual. Arrow indicates the location of the cortical break.

Five and three additional joints were excluded from JSW and Tt.BMD analyses at follow-up, respectively. There were no significant group-level changes in JSW, Tt.BMD or erosion volume over time at the 2^nd^ or 3^rd^ MCP (p > 0.05, Figure 4, Supplementary Figure 1). At the individual level, most joints did not exceed the LSC. However, four joints had decreased mean JSW and JSV, two had decreased BMD and four an increased erosion volume. Five joints had increased mean JSW, four had increased JSV, two had increased BMD and 2 had decreased erosion volume (Figure 4, Figure 5).

**Figure 4.**
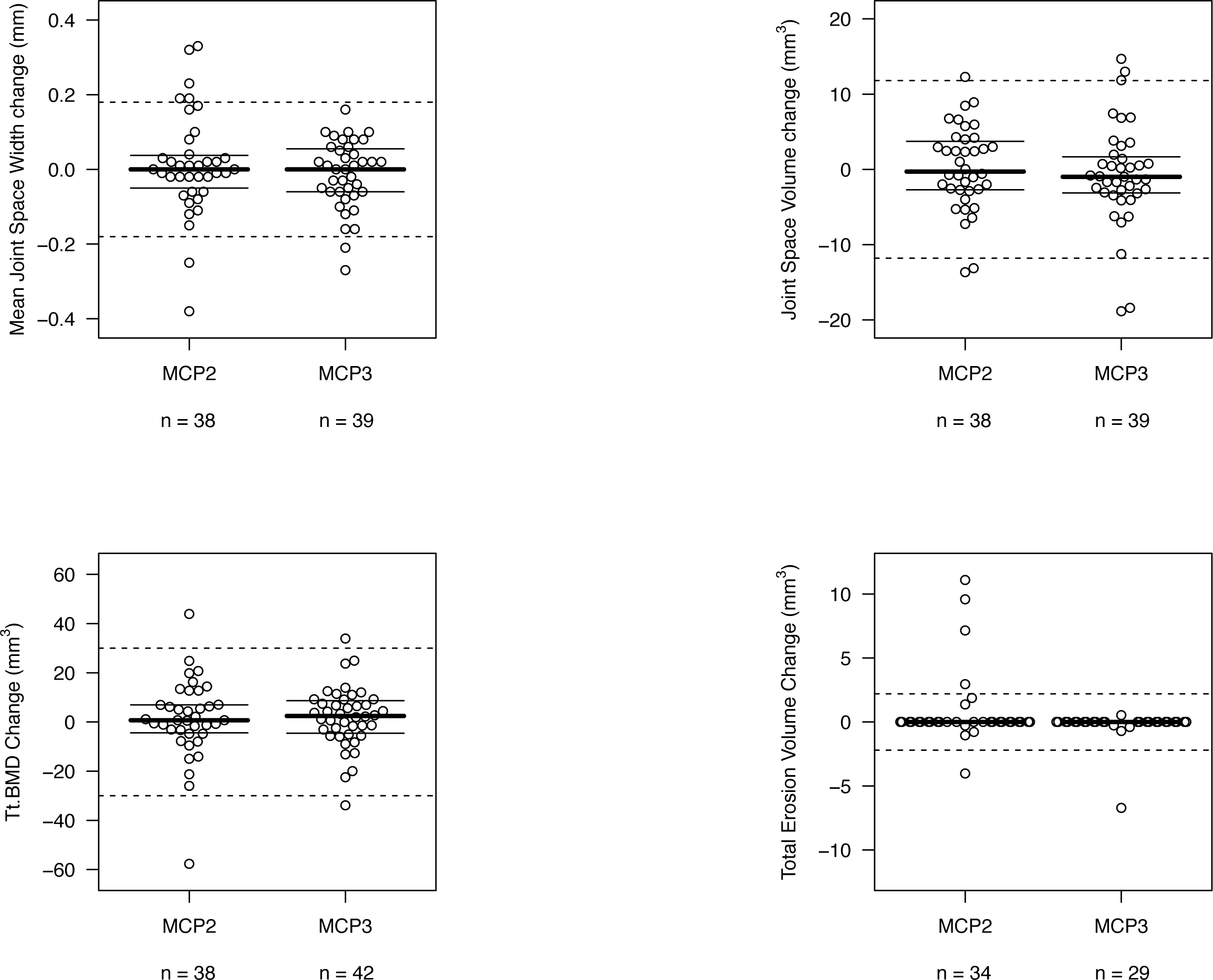
12-month changes in Mean JSW, JSV, TtBMD and Total Erosion Volume in the 2^nd^ MCP and 3^rd^ MCPs. Solid lines represent mean and inter-quartile range (IQR). Dashed lines represent least significant change (LSC).

**Figure 5.**
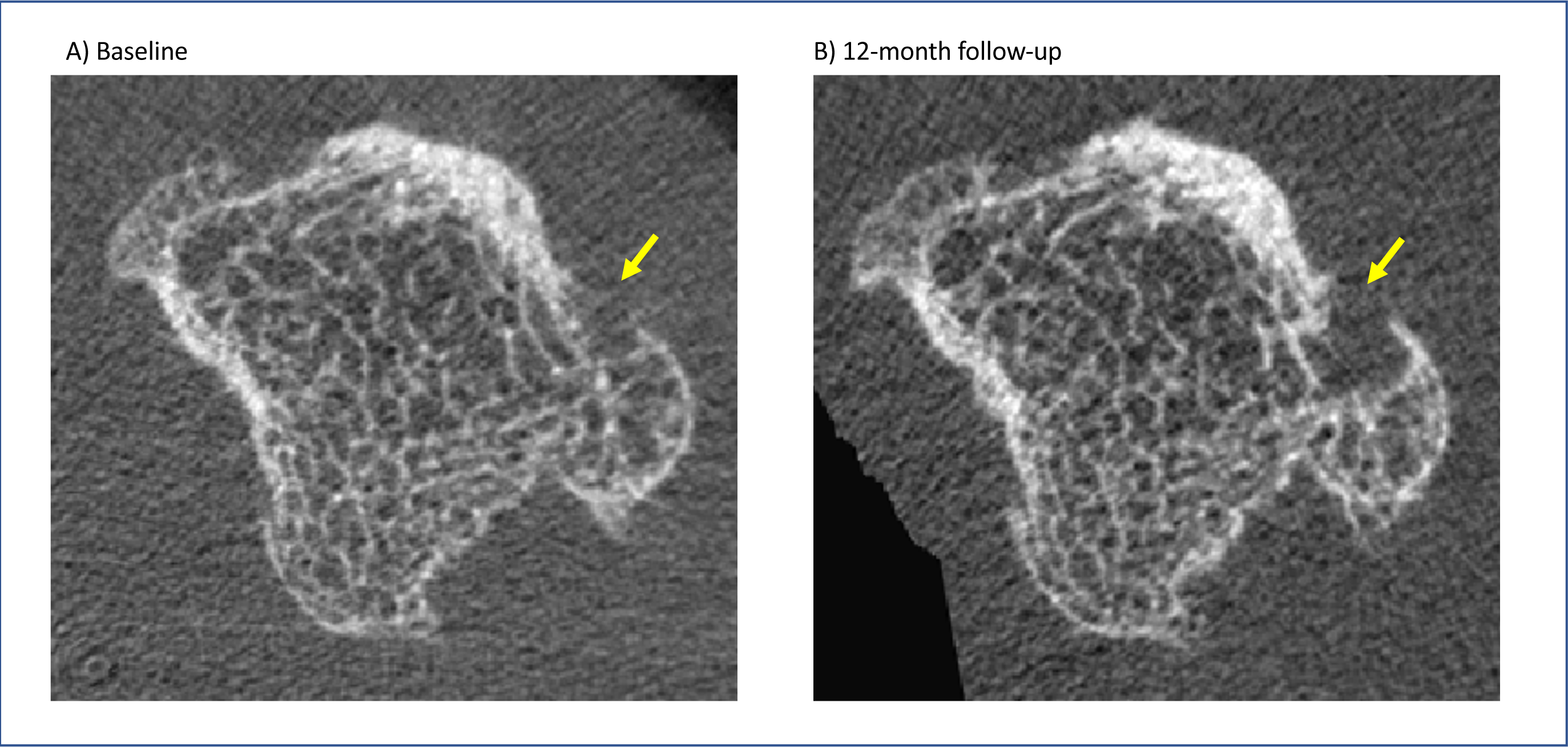
Axial HR-pQCT images of the 2^nd^ metacarpal demonstrating an erosion that increased in volume greater than the least significant change criterion. The 12-month follow-up image was registered and transformed to align with the baseline image to visually confirm the erosion change.

Patients who had a significant change in an HR-pQCT outcome were pooled into groups of improved (n = 7), stable (n = 16) or worsened (n = 9). No patient had both an improvement and worsening. Biologic DMARD-naïve patients were more likely to worsen, while bDMARD-treated patients were more likely to improve (p = 0.02). There was no difference in progress or improvement and disease activities at baseline or follow-up. Further, patients with undifferentiated or self-limited arthritis did not progress or improve differently from those with RA. Based on exploratory linear regression analyses, there were no other significant associations between baseline clinical measures and bone changes that were consistent among both the 2^nd^ and 3^rd^ MCPs.

### Radiographic Progression

Three patients increased in total vdHSS over 12-months. Of these patients, one had no HR-pQCT erosions but joint space progression on CR, one had no HR-pQCT erosions at baseline but developed one at follow-up, and one had erosions in the feet but not hands or wrists. No patient with only HR-pQCT-identified erosions showed an increase in vdHSS score.

## DISCUSSION

In this exploratory study in an EIA-cohort, HR-pQCT identified more erosions than CR in the specific joints examined, and could better characterize mimickers of erosions, such as pseudo-erosions. Despite clinical improvement over 12-months, we observed no group level changes in HR-pQCT-measured JSW, BMD or erosion volume, supporting that stability of disease both clinically and by imaging was common in our cohort.

While HR-pQCT is highly sensitive to changes in the joints we examined, the limited coverage presents challenges for full-patient assessment when compared with CR. Based on the few patients (3) who sustained radiographic progression, it is challenging to draw conclusions regarding the ability of HR-pQCT to predict radiographic progression. HR-pQCT is clearly best-suited to assessing erosion volume; the evaluation of the clinical relevance of detecting more erosions and changes in erosion volume in a small number of joints or detecting fewer erosions across a large number of joints will be the subject of future studies.

We sought to determine the relevance of pathology detected on HR-pQCT in early inflammatory arthritis. The correlation between minimum JSW and disease duration, while not strong, is consistent with findings of decreased radiographic JSN with disease duration.[38] Biologic DMARD-treated patients were more likely to demonstrate improvement in HR-pQCT outcomes, which is consistent with previous findings demonstrating decreases in erosion volumes under biologic therapies.[13,14,16] However, we did not observe any association with HR-pQCT outcomes and disease activity. The threshold for defining a significant change was based on precision of the outcomes at the 95% confidence level. We explored defining LSC at the 80% confidence level,[39] and although altering this threshold increased the number of patients experiencing a significant change, it did not reveal any additional associations with disease status. This suggests that determination of minimal clinically important differences for HR-pQCT is still required.

Our exploratory analysis found several prognostic factors associated with baseline HR-pQCT outcomes. The larger JSV, mean JSW and max JSW in males suggests that these outcomes are likely correlated with overall joint size. Therefore, sex and joint size may need to be considered in future HR-pQCT studies.

Our study has allowed us to demonstrate several technical advances in applying this imaging modality. We examined scan-rescan precision for HR-pQCT erosion and BMD analysis. Overall, the magnitude of the precision errors for erosion analysis, and poorer precision for scan-rescan than intra-rater analyses, reflect that the operator’s judgement is still required to delineate the surface of the erosion. This can be partly explained by using the first segmented image to guide the second analysis for scan-rescan analyses. In contrast, intra-rater analyses were performed without the guide of the previous erosion segmentation, three months apart. The poorer intra-rater precision in this study than previously reported by Töpfer et al. [9] may also have resulted from the time delay between intra-rater analyses, differences in analysis experience, and the larger mean erosion volume in our cohort (15.4 mm^3^ vs 9.3 mm^3^). The high RMSCV particularly for small erosions suggest that absolute precision errors should be reported alongside relative precision errors. And finally, with a greater number of erosions available for precision analysis, it would likely be prudent to calculate least-significant change values that are stratified by erosion size.

A limitation of the study was that there were a considerable number of observations that had to be discarded due to motion artifacts. Additionally, the discrepancy between new erosion identification from raw images to registered images suggests that registration of baseline to follow-up HR-pQCT images is critical to investigating subtle changes in erosion presence and volume.

In conclusion, HR-pQCT captures bone damage and progression undetectable by CR in the imaged joints. Incorporation of image registration and less operator-dependent erosion analyses will improve the ability to monitor change at the individual level.

Future studies of larger early inflammatory arthritis cohorts should investigate the minimal clinically important differences for HR-pQCT-assessed bone damage to determine what levels are associated with disease progression.

## Data Availability

Data will be made available upon reasonable request.

## ACKNOWLEDGEMENTS

We thank Tessa Linkert for assistance with patient recruitment and data collection, the staff and imaging technologists at the Centre for Mobility and Joint Health for data collection, as well as Sybren de Vries and Alicia Gabilondo for assistance with image analysis. We thank Dr. Charlie Goldsmith for assistance with statistical analysis. Study funding was provided by the University of Calgary. Scott Brunet was supported by the Natural Sciences and Engineering Research Council of Canada.

## FIGURE LEGENDS

Supplementary Figure 1. 12-month changes in minimum, maximum, SD and asymmetry of JSW. Solid lines represent mean and inter-quartile range (IQR). Dashed lines represent least significant change (LSC).

## REFERENCES

1 Cush JJ. Early rheumatoid arthritis -- is there a window of opportunity? J Rheumatol Suppl 2007;80:1–7.

2 Lard LR, Visser H, Speyer I, et al. Early versus delayed treatment in patients with recent-onset rheumatoid arthritis: comparison of two cohorts who received different treatment strategies. Am J Med 2001;111:446–51.

3 Korpela M, Laasonen L, Hannonen P, et al. Retardation of joint damage in patients with early rheumatoid arthritis by initial aggressive treatment with disease-modifying antirheumatic drugs: five-year experience from the FIN-RACo study. 2004;50:2072–81. doi:10.1002/art.20351

4 Manske SL, Zhu Y, Sandino C, et al. Human trabecular bone microarchitecture can be assessed independently of density with second generation HR-pQCT. Bone 2015;79:213–21. doi:10.1016/j.bone.2015.06.006

5 Boutroy S, Bouxsein ML, Munoz F, et al. In vivo assessment of trabecular bone microarchitecture by high-resolution peripheral quantitative computed tomography. J Clin Endocrinol Metab 2005;90:6508–15. doi:10.1210/jc.2005-1258

6 Burghardt AJ, Lee CH, Kuo D, et al. Quantitative in vivo HR-pQCT imaging of 3D wrist and metacarpophalangeal joint space width in rheumatoid arthritis. 2013;41:2553–64. doi:10.1007/s10439-013-0871-x

7 Barnabe C, Szabo E, Martin L, et al. Quantification of small joint space width, periarticular bone microstructure and erosions using high-resolution peripheral quantitative computed tomography in rheumatoid arthritis. Clin Exp Rheumatol 2013;31:243–50.

8 Peters M, Scharmga A, van Tubergen A, et al. The Reliability of a Semi-automated Algorithm for Detection of Cortical Interruptions in Finger Joints on High Resolution CT Compared to MicroCT. Calcif Tissue Int 2017;:1–9. doi:10.1007/s00223-017-0264-5

9 Töpfer D, Finzel S, Museyko O, et al. Segmentation and quantification of bone erosions in high-resolution peripheral quantitative computed tomography datasets of the metacarpophalangeal joints of patients with rheumatoid arthritis. Rheumatology (Oxford) 2014;53:65–71. doi:10.1093/rheumatology/ket259

10 Manske SL, Brunet SC, Finzel S, et al. The SPECTRA Collaboration OMERACT Working Group: Criterion Validity of Joint Space Outcomes with High Resolution Peripheral Quantitative Computed Tomography. J Rheumatol 2019;:jrheum.180870. doi:10.3899/jrheum.180870

11 Barnabe C, Sun Y, Boire G, et al. Heterogeneous Disease Trajectories Explain Variable Radiographic, Function and Quality of Life Outcomes in the Canadian Early Arthritis Cohort (CATCH). PLoS ONE 2015;10:e0135327. doi:10.1371/journal.pone.0135327

12 Skapenko A, Prots I, Schulze-Koops H. Prognostic factors in rheumatoid arthritis in the era of biologic agents. Nat Rev Rheumatol 2009;5:491–6. doi:10.1038/nrrheum.2009.157

13 Finzel S, Rech J, Schmidt S, et al. Repair of bone erosions in rheumatoid arthritis treated with tumour necrosis factor inhibitors is based on bone apposition at the base of the erosion. Ann Rheum Dis 2011;70:1587–93. doi:10.1136/ard.2010.148395

14 Shimizu T, Choi HJ, Heilmeier U, et al. Assessment of 3-month changes in bone microstructure under anti-TNFα therapy in patients with rheumatoid arthritis using high-resolution peripheral quantitative computed tomography (HR-pQCT). Arthritis Res Ther 2017;19:222. doi:10.1186/s13075-017-1430-x

15 Yue J, Griffith JF, Xiao F, et al. Repair of Bone Erosion in Rheumatoid Arthritis by Denosumab: A High-Resolution Peripheral Quantitative Computed Tomography Study. Arthritis Care Res (Hoboken) 2017;69:1156–63. doi:10.1002/acr.23133

16 Finzel S, Rech J, Schmidt S, et al. Interleukin-6 receptor blockade induces limited repair of bone erosions in rheumatoid arthritis: a micro CT study. Ann Rheum Dis 2013;72:396–400. doi:10.1136/annrheumdis-2011-201075

17 Fautrel B, Granger B, Combe B, et al. Matrix to predict rapid radiographic progression of early rheumatoid arthritis patients from the community treated with methotrexate or leflunomide: results from the ESPOIR cohort. Arthritis Res Ther 2012;14:R249. doi:10.1186/ar4092

18 Visser K, Goekoop-Ruiterman YPM, de Vries-Bouwstra JK, et al. A matrix risk model for the prediction of rapid radiographic progression in patients with rheumatoid arthritis receiving different dynamic treatment strategies: post hoc analyses from the BeSt study. Ann Rheum Dis 2010;69:1333–7. doi:10.1136/ard.2009.121160

19 Machold KP, Stamm TA, Nell VPK, et al. Very recent onset rheumatoid arthritis: clinical and serological patient characteristics associated with radiographic progression over the first years of disease. Rheumatology (Oxford) 2007;46:342–9. doi:10.1093/rheumatology/kel237

20 Dixey J, Solymossy C, Young A, et al. Is it possible to predict radiological damage in early rheumatoid arthritis (RA)? A report on the occurrence, progression, and prognostic factors of radiological erosions over the first 3 years in 866 patients from the Early RA Study (ERAS). J Rheumatol Suppl 2004;69:48–54.

21 Aletaha D, Neogi T, Silman AJ, et al. 2010 Rheumatoid arthritis classification criteria: an American College of Rheumatology/European League Against Rheumatism collaborative initiative. 2010;62:2569–81. doi:10.1002/art.27584

22 Bruce B, Fries JF. The Health Assessment Questionnaire (HAQ). Clin Exp Rheumatol 2005;23:S14–8.

23 Wells G, Becker J-C, Teng J, et al. Validation of the 28-joint Disease Activity Score (DAS28) and European League Against Rheumatism response criteria based on C-reactive protein against disease progression in patients with rheumatoid arthritis, and comparison with the DAS28 based on erythrocyte sedimentation rate. Ann Rheum Dis 2009;68:954–60. doi:10.1136/ard.2007.084459

24 Fransen J, van Riel PLCM. DAS remission cut points. Clin Exp Rheumatol 2006;24:S– 29–32.

25 van der Heijde D, Paulus H, Shekelle P. How to read radiographs according to the Sharp/van der Heijde method. Discussion : Heterogeneity in rheumatoid arthritis radiographic trials. Issues to consider in a metaanalysis. J Rheumatol 2000;27:261–3.

26 Krabben A, Stomp W, van Nies JAB, et al. MRI-detected subclinical joint inflammation is associated with radiographic progression. Ann Rheum Dis 2014;73:2034–7. doi:10.1136/annrheumdis-2014-205208

27 Barnabe C, Buie H, Kan M, et al. Reproducible metacarpal joint space width measurements using 3D analysis of images acquired with high-resolution peripheral quantitative computed tomography. Med Eng Phys 2013;35:1540–4. doi:10.1016/j.medengphy.2013.04.003

28 Barnabe C, Feehan L, SPECTRA (Study GrouP for XTrEme-CT in RA). High-resolution peripheral quantitative computed tomography imaging protocol for metacarpophalangeal joints in inflammatory arthritis: the SPECTRA collaboration. J Rheumatol 2012;39:1494–5. doi:10.3899/jrheum.120218

29 Stok KS, Burghardt AJ, Boutroy S, et al. Consensus approach for 3D joint space width of metacarpophalangeal joints of rheumatoid arthritis patients using high-resolution peripheral quantitative computed tomography. Quant Imaging Med Surg

30 International Society for Clinical Densitometry Committee on Standards of Bone Measurement. 2015 ISCD Official Positions - Adult. Published Online First: 2015.http://www.iscd.org/official-positions/2015-iscd-official-positions-adult/

31 Glüer CC, Blake G, Lu Y, et al. Accurate assessment of precision errors: how to measure the reproducibility of bone densitometry techniques. Osteoporos Int 1995;5:262–70.

32 Glüer CC. Monitoring skeletal changes by radiological techniques. J Bone Miner Res 1999;14:1952–62. doi:10.1359/jbmr.1999.14.11.1952

33 Eklund A. beeswarm. http://www.cbs.dtu.dkeklundbeeswarm.

34 Prevoo ML, van ’t Hof MA, Kuper HH, et al. Modified disease activity scores that include twenty-eight-joint counts. Development and validation in a prospective longitudinal study of patients with rheumatoid arthritis. 1995;38:44–8.

35 Team RS. RStudio: Integrated Development for R. RStudio, Inc. 2015.http://www.rstudio.com/ (accessed 26 Apr2019).

36 Allaire JJ, Xie Y, McPherson J, et al. rmarkdown: Dynamic Documents for R. R package version 1.12. 2019.https://rmarkdown.rstudio.com (accessed 26 Apr2019).

37 Xie Y, Allaire JJ, Grolemund G. R Markdown: The Definitive Guide. Chapman and Hall/CRC 2018. https://bookdown.org/yihui/rmarkdown

38 Fuchs HA, Kaye JJ, Callahan LF, et al. Evidence of significant radiographic damage in rheumatoid arthritis within the first 2 years of disease. J Rheumatol 1989;16:585–91.

39 Navarro-Compán V, van der Heijde D, Ahmad HA, et al. Measurement error in the assessment of radiographic progression in rheumatoid arthritis (RA) clinical trials: the smallest detectable change (SDC) revisited. Ann Rheum Dis 2014;73:1067–70. doi:10.1136/annrheumdis-2012-202939

